# Violent and Nonviolent Death Tolls for the Gaza War: New Primary Evidence

**DOI:** 10.1101/2025.06.19.25329797

**Authors:** Michael Spagat, Jon Pedersen, Khalil Shikaki, Michael Robbins, Eran Bendavid, Håvard Hegre, Debarati Guha-Sapir

## Abstract

High-quality war mortality estimates, such as those that were produced for Kosovo, Iraq, and Darfur play a critical role in illuminating the human cost of war. During the tumult of war they are, however, challenging to obtain due to the conflicts themselves. The Gaza Ministry of Health (GMoH) has provided regular updates to their post-October-7 violent death tally for the Gaza Strip. However, GMoH reports have attracted both criticism and support. Here, we present results from a large-scale household survey, the Gaza Mortality Survey (GMS), which provides independent estimates of war-related deaths between October 7, 2023 and January 5, 2025. Our findings suggest that violent mortality has significantly exceeded official figures. Our central estimate for the extent of GMoH undercount closely matches a separate estimate made using capture-recapture methods. We also find that nonviolent excess deaths, often overlooked in conflict assessments, also represent a substantial burden. These results underscore the feasibility of mortality surveillance methodologies in some highly challenging war-affected settings and provide a crucial empirical foundation for assessing the true human cost of the war.

## 1. INTRODUCTION

High-quality war mortality estimates, such as those that were produced for Kosovo, Iraq, and Darfur (1–3) play a critical role in illuminating the human cost of war. During the tumult of war they are, however, challenging to obtain due to the conflicts themselves. The Gaza Ministry of Health (GMoH) has provided regular updates to their post-October-7 violent death tally for the Gaza Strip. However, GMoH reports have attracted both criticism (4–5) and support (6–8). Here, we present results from a large-scale household survey, the Gaza Mortality Survey (GMS), which provides independent estimates of war-related deaths between October 7, 2023 and January 5, 2025. Our findings suggest that violent mortality has significantly exceeded official figures. Our central estimate for the extent of GMoH undercount closely matches a separate estimate made using capture-recapture methods (7). We also find that nonviolent excess deaths, often overlooked in conflict assessments, also represent a substantial burden. These results underscore the feasibility of mortality surveillance methodologies in some highly challenging war-affected settings and provide a crucial empirical foundation for assessing the true human cost of the war.

A recent study triangulating 2 sources of GMoH data with social media obituaries (7) yielded a violent death toll estimate of 64,260 deaths (95% CI 55,298–78,525) from the beginning of the war through June 15, 2024. This work makes substantive contributions to generating independent estimates of the Gaza death toll, yet (7) has meaningful limitations. First, two of their three data sources, and a strong majority of the deaths, are components of the GMoH database itself. So (7) can offer only limited and partially circular assessment of GMoH overall numbers and their demographic disaggregates. Second, their approach – a three-list capture recapture – admits multiple valid models that are all plausible given our current state of knowledge. As a group, the 95% confidence intervals for these models range from 47,457 to 88,332 deaths. So, even if we accept the broad thrust of the analysis in (7), considerable uncertainty remains (9), and fully independent assessment of the death toll – one not reliant on GMoH data – is needed to clarify the violent death toll of the war. Finally, major studies that estimate the death toll in Gaza based on GMoH data (7–8) fail to shed light on the number of nonviolent excess deaths caused indirectly by the war.

Indeed, information on the nonviolent excess death toll of the war is, to date, scant and unreliable. A report on this topic (10) estimated that it that the number of nonviolent deaths in the war could be 4 times as high as the number of violent deaths. Additionally, physicians who worked in the Gaza Strip (11) claimed tens of thousands of deaths due to starvation. However, these studies are not based on systematic data collection. To date, no empirically grounded estimates of nonviolent deaths for the Gaza war exist, a gap we aim to fill.

In this study, the Gaza Mortality Survey (GMS), we present newly and independently collected survey data from the Gaza Strip that enable fresh and more precise estimates of both the violent and nonviolent death toll of the war. In the GMS, we surveyed a representative sample of 2,000 Gazan households (representative of Gaza pre-Pre-October 7) and collected information on the vital status of 9,729 household members and their newborns, including whether they are alive or dead. Reported deaths are divided into violent or nonviolent ones.

Based on the GMS, we estimate 75,200 violent deaths (95% CI: 63,600– 86,800) between October 7, 2023, and January 5, 2025. Among those, 56.2% (95% CI: 50.4%–61.9%) were women (18–64), children (< 18), or elderly (65+). We also estimate 8.540 (95% CI: 4,540–12,500) excess (i.e., above expectation) nonviolent deaths.

The comparable, i.e., for the same period, GMoH estimate of about 49,090^1^ is roughly 34.7% below our central estimate and 22.8% below the lower bound of our 95% confidence interval for violent deaths, suggesting that the GMoH has not exaggerated the death toll in the war. Our estimated percentage of women, children, and elderly among the dead is consistent with GMoH reporting that put this number at 54.0% for the GMS coverage period (GMoH data). We also provide grounded estimates for the number of excess nonviolent deaths, an original contribution.

These results demonstrate the feasibility of data collection, even in environments as challenging as the Gaza Strip during the present war and highlight the need for improved mortality surveillance in conflict zones.

## 2. COLLECTION AND DESCRIPTION OF THE SAMPLE

### 2.1 Sampling and Fieldwork

The Materials and Methods file of the supplementary materials section provides more details on GMS sampling and fieldwork than this brief overview does and still more details are in Supplementary Information (SI).

The survey was preregistered, ethically approved by Royal Holloway University of London and obtained informed consent from each respondent. The Palestinian Center for Policy and Survey Research (PCPSR) collected the data from 2,000 households spread across 200 Primary Sampling Units (PSUs). Challenges to drawing a representative sample included widespread displacement and the inaccessibility of Northern Gaza, Gaza City, and Rafah due to ongoing fighting and Israeli evacuation orders. PCPSR, which has tracked population movements throughout the war, randomly selected 70, 41 and 89 PSUs in, respectively, census enumeration areas, built-up shelters, and tent gatherings - mirroring the population proportions by dwelling type when the survey was conducted. We strived to ensure representativeness by governorate of origin by earmarking some shelter and tent PSUs to cover the populations of the three inaccessible governorates. Figure 1 shows PSU locations and the governorate of origin that each PSU represents. We describe the selection procedures within PSUs in the SI section.

**Figure 1:**
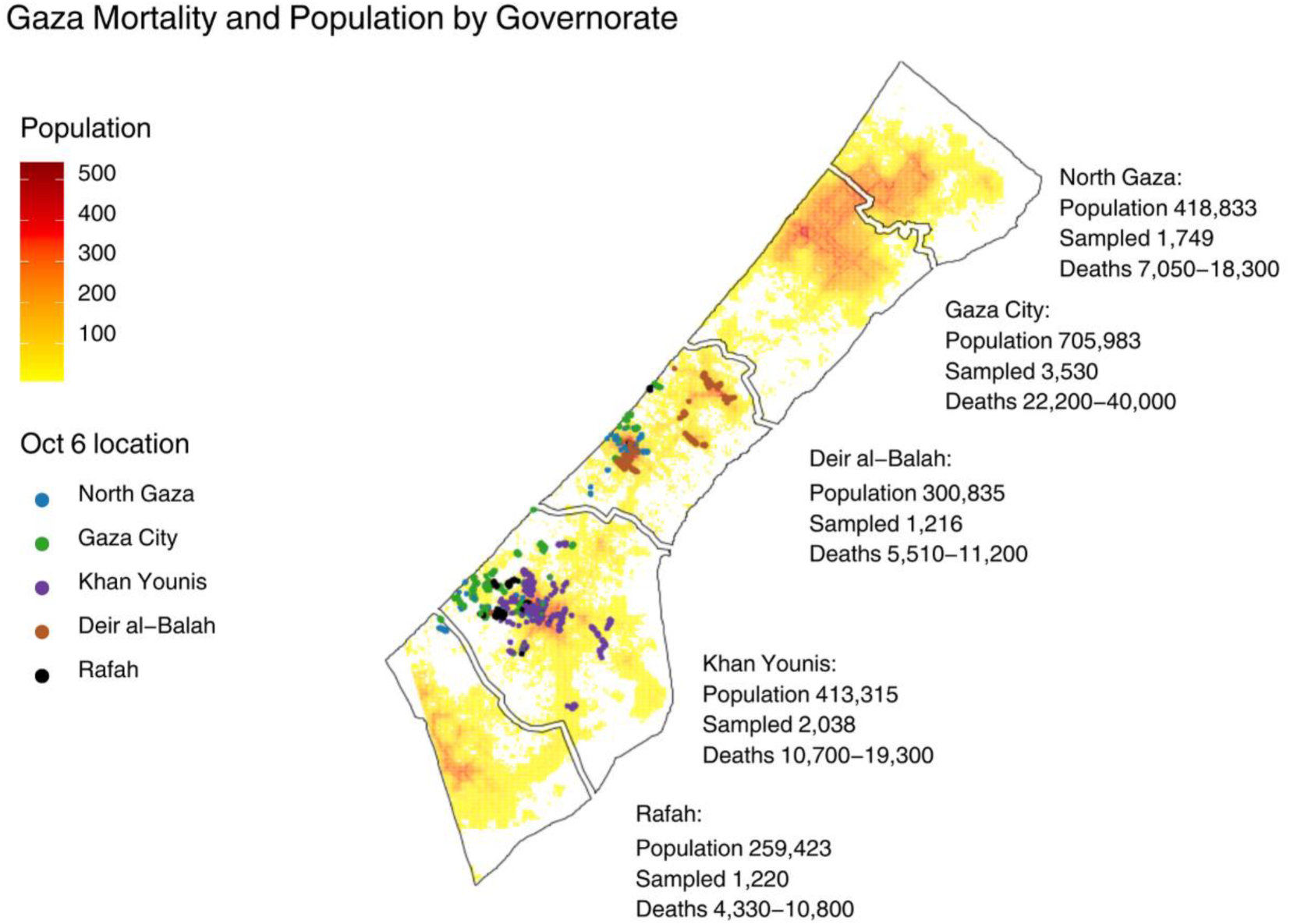
Sampling points by location, colour coded for governorate of origin, of respondents with further information on the sample and estimates.

We wrote the questionnaire in English and PCPSR translated and then tested it in Arabic. The interview documents household members as of October 6, 2023, plus newborns added to these households during the war, and their post-October-7 vital status. Interviewers were trained to ensure accurate mortality reporting, focusing only on household members. Data were collected between December 30, 2024, and January 5, 2025, by ten two-person teams supported by four supervisors and other PCPSR staff. GPS tracking and real-time monitoring were among the quality control measures. Face-to-face interviews were conducted using tablets and mobile phones, with data instantly uploaded to a secure central server.

### 2.2. Sample Description

Table 1 shows the status, by gender and age, of all people recorded by the survey as household members as of October 6, 2023. Between the ages of 5 and 39, deaths among men greatly outnumber deaths among women. There are more men than women reported missing in all age categories, including children, especially for ages 18-39. Women make up just 1 out of 52 people reported imprisoned. The primary group in the sample who left Gaza are young adults.

**Table 1:** Outcomes for all people in the sample, classified by gender and age.

| Gender/Age | Present | Left Gaza | Moved Out of Household | Dead | Missing | In Prison | Total |
| --- | --- | --- | --- | --- | --- | --- | --- |
| Male 0-4 | 509 | 5 | 0 | 8 | 1 | 0 | 523 |
| Female 0-4 | 441 | 1 | 0 | 10 | 0 | 0 | 452 |
| Male 5-11 | 757 | 12 | 3 | 27 | 2 | 0 | 801 |
| Female 5-11 | 679 | 13 | 4 | 17 | 2 | 0 | 715 |
| Male 12-17 | 534 | 11 | 2 | 31 | 7 | 0 | 585 |
| Female 12-17 | 581 | 13 | 3 | 12 | 2 | 0 | 611 |
| Male 18-29 | 1015 | 65 | 30 | 106 | 27 | 21 | 1264 |
| Female 18-29 | 1277 | 53 | 33 | 53 | 0 | 0 | 1416 |
| Male 30-39 | 631 | 24 | 20 | 52 | 14 | 23 | 764 |
| Female 30-39 | 635 | 30 | 6 | 20 | 0 | 0 | 691 |
| Male 40-64 | 817 | 22 | 24 | 42 | 6 | 6 | 917 |
| Female 40-64 | 721 | 28 | 8 | 43 | 1 | 1 | 802 |
| Male 65+ | 81 | 0 | 2 | 17 | 0 | 1 | 101 |
| Female 65+ | 62 | 4 | 2 | 19 | 0 | 0 | 87 |
| Total | 8740 | 281 | 137 | 457 | 62 | 52 | 9729 |
| Unweighted GMS Data |  |  |  |  |  |  |  |

Table 2 subdivides reported deaths into categories, by gender and age. Violent deaths greatly outnumber nonviolent deaths. Among violent deaths, there are many more males than females, especially between the ages of 18-39.

**Table 2:** All deaths in the sample, classified by gender and age.

| Gender/Age | Nonviolent | Accident | Violent | Total |
| --- | --- | --- | --- | --- |
| Male 0-4 | 0 | 0 | 8 | 8 |
| Female 0-4 | 2 | 0 | 8 | 10 |
| Male 5-11 | 2 | 2 | 23 | 27 |
| Female 5-11 | 0 | 1 | 16 | 17 |
| Male 12-17 | 0 | 0 | 31 | 31 |
| Female 12-17 | 1 | 0 | 11 | 12 |
| Male 18-29 | 2 | 0 | 104 | 106 |
| Female 18-29 | 0 | 0 | 53 | 53 |
| Male 30-39 | 0 | 1 | 51 | 52 |
| Female 30-39 | 1 | 0 | 19 | 20 |
| Male 40-64 | 8 | 0 | 34 | 42 |
| Female 40-64 | 17 | 0 | 26 | 43 |
| Male 65+ | 9 | 1 | 7 | 17 |
| Female 65+ | 17 | 0 | 2 | 19 |
| Total | 59 | 5 | 393 | 457 |
| Unweighted GMS Data |  |  |  |  |

Table 3 gives the within-sample number of violent deaths along with the percentage dying violently within each governorate. Readers should interpret this table with caution because many people, after being displaced, will have died outside their governorate of origin, and we did not collect information on where people were killed.

**Table 3:** All violent deaths in the sample, classified by governorate of origin.

| Governorates | Violent Deaths | Violent Deaths as Percent of Sample |
| --- | --- | --- |
| Northern Gaza | 68 | 3.9 |
| Gaza City | 164 | 4.7 |
| Deir al-Balah | 39 | 3.2 |
| Khan Younis | 87 | 4.3 |
| Rafah | 35 | 2.9 |
| Unweighted GMS Data |  |  |

The survey recorded post-October-6 births of 179 boys and 178 girls, of whom it was reported that two boys and two girls had died.

## 3. ESTIMATES

To adjust for the sample not corresponding exactly to expected distributions of the population by ages, gender, regions in the Gaza Strip (governorates), and household sizes, we raked (13–14) the sample with marginal distributions derived from the US Census Bureau population projections for ages and genders (15), the Palestinian Central Bureau of Statistics for their projections of population by governorate in the Gaza Strip (16), and the Multiple Indicator Cluster Survey 2019-20 (17) for the distribution of household sizes. We trimmed high weights down to twice the median weight for the whole sample, performing all the work with the survey.design and trimWeights procedure of the R package Survey 4.4.2 (18). Table S6 in supplementary materials shows the impact of raking and trimming on the estimates. We also used the linearization method in Survey to estimate standard errors.

Table 4 displays our main estimates. We stress ranges over central estimates and caution readers that, as with any survey, we cannot quantify all the uncertainty affecting the estimates.

**Table 4:**
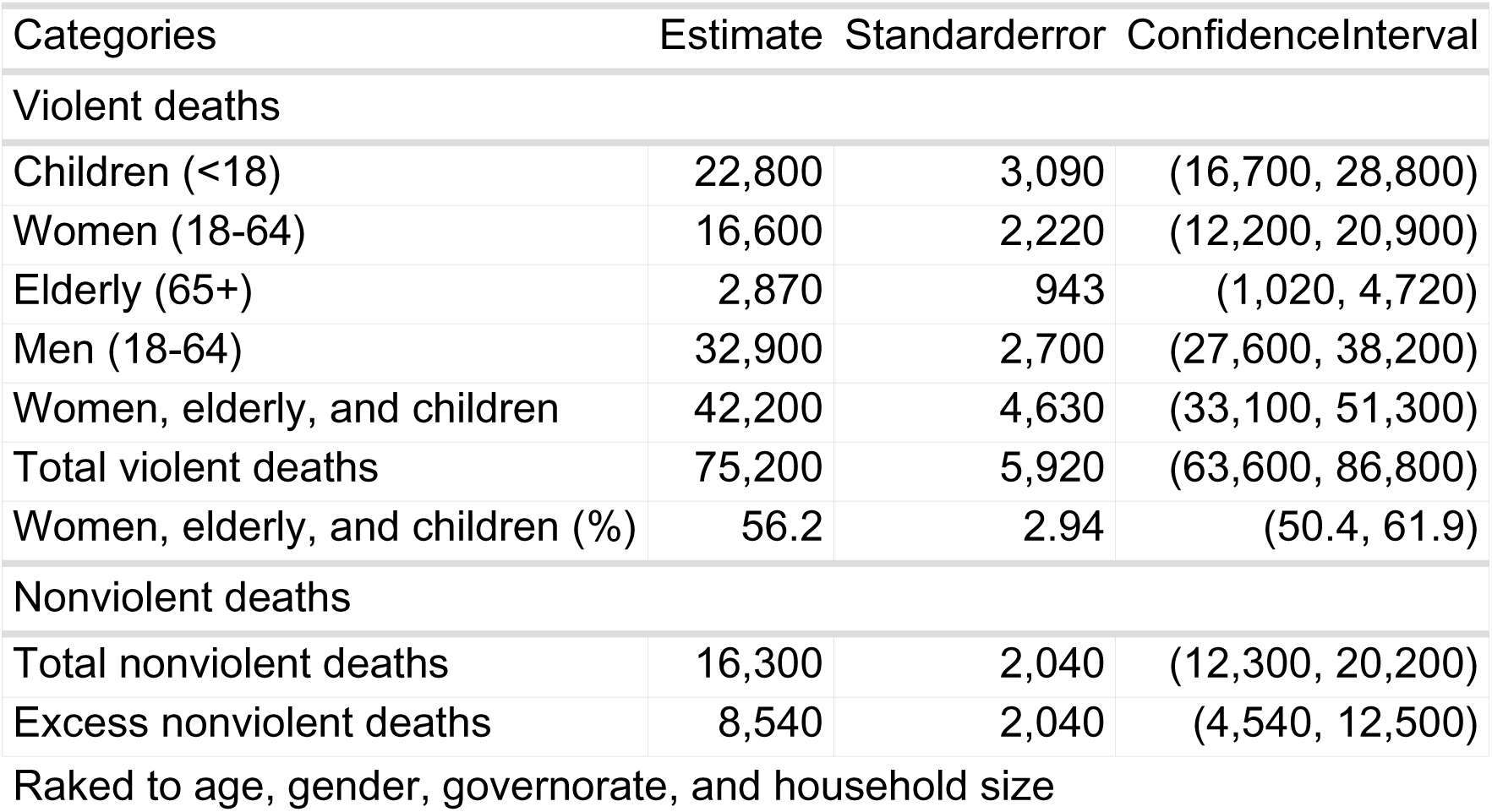
Estimates of violent and nonviolent (including accidents) mortality by broad age and gender groups.

| Categories | Estimate | Standarderror | ConfidenceInterval |
| --- | --- | --- | --- |
| Violent deaths |  |  |  |
| Children (<18) | 22,800 | 3,090 | (16,700, 28,800) |
| Women (18-64) | 16,600 | 2,220 | (12,200, 20,900) |
| Elderly (65+) | 2,870 | 943 | (1,020, 4,720) |
| Men (18-64) | 32,900 | 2,700 | (27,600, 38,200) |
| Women, elderly, and children | 42,200 | 4,630 | (33,100, 51,300) |
| Total violent deaths | 75,200 | 5,920 | (63,600, 86,800) |
| Women, elderly, and children (%) | 56.2 | 2.94 | (50.4, 61.9) |
| Nonviolent deaths |  |  |  |
| Total nonviolent deaths | 16,300 | 2,040 | (12,300, 20,200) |
| Excess nonviolent deaths | 8,540 | 2,040 | (4,540, 12,500) |

Raked to age, gender, governorate, and household size

The range of our violent death estimate, 63,600–86,800, is between 3.0-4.1% of the population of the Gaza Strip. The GMoH figure of 45,650 is 53% of the top of our 95% confidence interval for violent deaths and 72% of the bottom of this interval.

We estimate that women, children (<18) and elderly (65+) make up 56.2% (95% CI: 50.4%–61.9%) of the violent deaths during the study period, very close to the comparable GMoH percentage of 54.0% (GMoH data). In absolute terms, the bottom of our 95% confidence intervals for this group (33,100) exceeds the comparable GMoH figure of 26,509 (GMoH data).

We estimate a further 16,300 (95% CI 12,300-20,200) nonviolent deaths, the first such estimate for the Gaza Strip during the war. To estimate excess nonviolent deaths, we subtract 7,716 deaths predicted (19) to occur without war, a number that combines projections of 6,348 deaths for 2023 and 6,188 deaths for 2024 and 6,043 deaths for 2025, prorated for the number of war days within each year. This procedure produces an excess nonviolent death estimate of 8,540 (95% CI: 4,540– 12,500), in line with an early projection for a scenario under which violence escalates but epidemics are avoided of 5,780 covering February through August 2024 (20).

## 4. DISCUSSION

### 4.1 Interpretations and Conclusion

In this survey-based study of mortality in the Gaza war through January 5 2025, we conclude that: (a) the violent death toll of the war resulted in the deaths of around 3.6% of Gaza’s population, exceeding the official GMoH total; (b) around 56% of the violent deaths were among women, children, and the elderly, a percentage similar to that endorsed by the GMoH; and (c) violent deaths far outnumber nonviolent deaths, though there have still been at least several thousand nonviolent excess deaths. The proportion of deaths made up by women, children, and elderly supports the general perception in which non-combatants are frequent victims of the conflict.

There is striking alignment between our survey-based finding, that the comparable GMoH violent-death number is 34.7% below our central estimate, and the finding of (7), using completely different capture-recapture methods, that the GMoH tally is 39.1% below their central estimate.^2^ Thus, the GMO and (7) combined provide a rare validation (21) finding for the two methodologies which are increasingly used to measure death tolls in war (22, 23).

Our findings are also incompatible with claims that: (a) the GMoH has “inflated the death toll” (5, 24); (b) indirect deaths could exceed violent deaths by at least a factor of four (10, 25); and (c) “it is likely that 62,413 people have died of starvation” (appendix to 11). Follow-up surveys might provide central estimates or distributions that differ substantially from the GMS. Nevertheless, this is the first population survey of mortality in Gaza which gives independent and grounded estimates that will anchor future discussions.

Our findings suggests that a high ratio of indirect to direct deaths is not inevitable in warfare, as some analysts seem to assume (10). This should not be surprising as, for example, the Kosovo Memory Book project classified just 281 out of 13,517 deaths in the Kosovo war (1998–1999) as indirect (26). Indeed, indirect deaths are usually only estimated (27) in settings with extremely poor conditions such as Darfur (3) or Tigray (28) and direct deaths have substantially outnumbered indirect ones even in the extremely impoverished environment of Yemen (26).

Despite a claim to the contrary (7), the GMS demonstrates that a ground survey in the Gaza Strip was feasible despite the challenging conditions. This success highlights the need for improved mortality surveillance in conflict zones.

The GMoH work has value that extends far beyond the insight it provides into the total number of violent deaths in the war and the number’s demographic composition. By naming individual victims one by one, the GMoH endows each person with a measure of human dignity (27). This project of memorialization has only begun and must continue for many years after the war ends. Estimates of numbers killed, the focus of the present paper, can help guide this casualty recording work. But we cannot provide each human being due recognition in death simply by estimating the number of deaths.

### 4.2 Sources of bias and uncertainty

Here we list and evaluate potential sources of both upward and downward bias in our estimates, beginning with upward bias.

Respondents may have overreported deaths, perhaps with the objective of collecting compensation for dead relatives or otherwise inflating the conflict’s mortality counts. Interviewers were clear, however, that respondents could not gain materially from their participation. Moreover, interviews began with a household roster before ascertaining the fate of each person listed on the roster. This procedure was intended to limit inflating death counts by forcing respondents to report deaths only of previously enumerated individuals, without *post hoc* addition of deaths.

Respondents may have reported deaths that are real but of relatives who were not household members, as seems to have happened in a survey of Iraq (28). We anticipated this possibility. Interviewer training stressed this and emphasized that relatives who are not household members must not be included in the household roster. Indeed, the mean household size in the sample (4.9) was below the population figure (5.5), arguing against inflation of households with non-member relatives.

We could not sample some populations. For example, we could not reach households that had left the Gaza Strip entirely (i.e., all members left). Although some people have left to receive medical treatment, or because of deaths of household members, it is plausible that leaving Gaza Strip is correlated with lower mortality, both because of better conditions in the destination place of residence, as well as because of selection (families with more means may have been more likely to leave). We note, however, that the number of leavers identified in our sample was relatively small (Table S4).

Finally, survey interview teams may have preferentially selected relatively high-mortality households. The survey protocol they were instructed to follow is designed to prevent this from happening, since we recognize that inadvertent factors may drive sympathetic human interviewers towards more affected families. The extent to which this may have happened is hard to ascertain and we have no positive evidence that it did happen. However, we address the possibility through a scenario in our sensitivity analysis (Table S6) that removes the data collected by the survey team that captured the most violent deaths.

A source of downward bias is our non-sampling of households with zero remaining live members, or no live members above the age of 15, whom we could interview. While the number of such households is unclear, this phenomenon has been documented by others (29).

We used shelter interviews exclusively to capture households from Northern Gaza, Gaza City and Rafah. Therefore, households from Deir al-Balah or Khan Younis that were displaced into shelters were outside our sampling frame. Thus, the estimates assume, implicitly, that mortality rates for displaced and nondisplaced households from these governorates were equal, although it seems likely that the former group suffered higher mortality than the latter group.

We were unable to reach households that remained entirely in Northern Gaza, Gaza City, or Rafah due to the extreme danger in these areas (30). This gap could contribute to underestimation. However, many displaced households experienced violence during departure, transit, or in their new locations. While we lack direct evidence on how these dynamics offset each other, the scale of displacement suggests that excluding those who remained is likely a minor source of bias.

It is unclear to what extent individuals classified as missing may be dead. This has implications not only for the overall mortality numbers, but also for the portion of the violent deaths made up by women, children, and elderly. Nevertheless, it is worth noting that, even if we make the extreme and implausible assumption that all missing persons are dead, then the central estimate for the percentage of women, children, and elderly among the dead decreases only to 52.2% (Table S5).

The above discussion of biases shows that, as is the case with all surveys, confidence intervals cannot capture all the important uncertainty surrounding the estimates. Moreover, we did not attempt to model uncertainty over the number of projected deaths in the absence of war, instead using a single number for baseline deaths in our excess death calculation while acknowledging that this is a further source of uncertainty. Readers should, therefore, view our confidence intervals as minimum ranges.

### 4.3 The Future

A fragile ceasefire began roughly 10 days after our data collection ended but collapsed with renewed Israeli airstrikes on March 18. The humanitarian situation has further deteriorated since then. Widespread destruction across the Gaza Strip continues to cripple institutions essential to human development, particularly the health care system. Undercounting of violent deaths by the GMoH is likely to persist, although not necessarily at the GMS-estimated rate. On April 7, six senior UN officials warned of a looming humanitarian catastrophe (31), and subsequent reports through May and early July indicate worsening conditions. The ratio of nonviolent to violent deaths may well have grown since the GMS data collection period. Past scholarship suggests that the long-term toll of conflict will probably accumulate over the coming years even if the acute conflict ends (35–37). This underscores the importance of continued GMoH surveillance, as well as independent surveys, to properly understand the unfolding dynamics of death. We hope that resources will be available to tackle this task.

## Data Availability

All data produced are available online at Github

https://gitfront.io/r/mspagat/tZwP79d7Pntz/Gaza-Mortality-Survey/

## Methods

The GMS is preregistered at OSF (1) and has ethical approval from Royal Holloway University of London. The Palestinian Center for Policy and Survey Research (PCPSR) collected the data. The rest of this section describes the sampling and field work. A companion document, also included in supplementary materials, provided further details.

### 1. SAMPLING

We sampled 2,000 households at 200 Primary Sampling Units (PSUs) with 10 interviews conducted at each PSU. In rare cases, PSU size differed slightly due to replacement interviews being collected in a different location after the team had moved on. The response rate was also extremely high, with only 58 eligible households refusing to be interviewed.

There were two main challenges to address in pursuit of the goal of collecting a representative sample for the whole Gaza Strip. First, most of the population was displaced and could not be located through the census. Second, active combat or Israeli evacuation orders rendered the governorates of Northern Gaza, Gaza City and Rafah inaccessible during the field work. However, for the sample to be representative, households originating in these hot zones needed to be included.

PCPSR coordinated with the local government authorities, UNRWA and other relevant international organizations to address the displacement challenge by tracking population movements throughout the war. This work resulted in the division of current living arrangements at the time of the survey into three types.^3^ Each of these residency types served as a primary stratum for the survey:

1) **Enumeration areas**

The Palestinian Central Bureau of Statistics (PCBS) has provided PCPSR with a sample of enumeration areas from the 2017 Palestinian Census, each containing on average 143 dwelling units. The sample includes enumeration area maps with all pre-war homes, streets and public places numbered and now also has a list of still-occupied enumeration areas, in Khan Younis and Deir al-Balah (the central Gaza Strip). The enumeration areas were selected with inclusion probabilities proportionate to their number of households.

2) **Built-up shelters**

These are buildings, mostly schools, universities, clubs and other standing structures that existed before the war and have since been converted into shelters. PCPSR has constructed a list of all the shelter centers in Deir al-Balah and Khan Younis governorates which it has divided into blocks of roughly equal population sizes. The blocks were selected with linear systematic simple random sampling.

3) **Tent gatherings**

These are shelters that were created after the war started in the areas of Khan Younis and Deir al-Balah governorates. PCPSR has satellite maps showing the areas covered by these communities which it has divided into blocks of roughly equal population sizes. Again, the blocks were selected with linear systematic random sampling.

At the moment of sample selection PCPSR estimated that roughly 35% of the population were in enumeration areas, 20.5% in built-up shelters and 44.5% in tent gatherings. Accordingly, our random selection of 200 PSU’s contains 70 occupied enumeration areas, 41 built shelter blocks and 89 blocks of tent gatherings.

Households were numbered within each selected PSU. Within PSUs that were enumeration areas the interview teams proceeded systematically from a randomly selected starting number through nine further households at a fixed interval with the goal of distributing the interviews across the entire PSU.

**Table M1.**
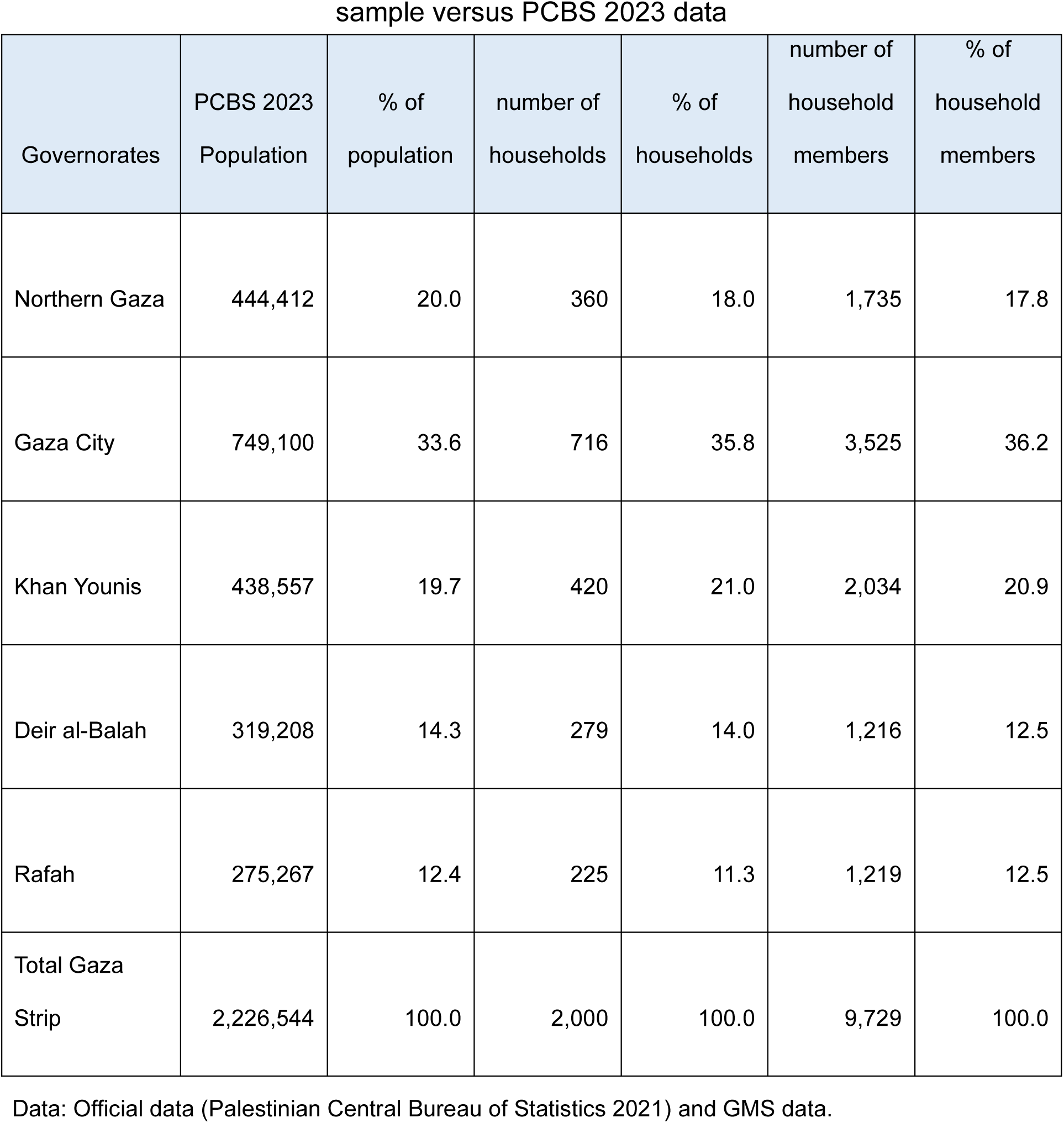
Households and household members by governorate: sample versus PCBS 2023 data.

To address the challenge, noted above, of including households from areas we could not access during the field work, the remaining 130 PSU’s, all in built-up shelters or tent gatherings, were sub-stratified by governorate of residency based on October 6, 2023, for inaccessible governorates. Accordingly, within built up shelters and tent shelters, each PSU was assigned to include only residents displaced from either Northern Gaza, Gaza City or Rafah. Thus, in a Northern-Gaza-designated PSU, for example, the interview team would first ask if a household had originated in Northern Gaza. If the answer was “no” then the team would immediately terminate the interview and proceed to the next household according to the fixed interval set for that PSU. The team would continue with these initial probes and actual interviews until they completed 10 interviews of households from Northern Gaza in that PSU. Table M1 suggests that these procedures did succeed in drawing a sample that closely mirrors the pre-war population distribution by governorate in the Gaza Strip.

In summary, the sample is a stratified (by governorate/dwelling type pairs) two-stage cluster sample.

## 2. FIELD WORK

We first developed a questionnaire in English that was then passed to a PCPSR team which recommended a few changes and translated it into Arabic appropriate for the Gaza Strip. PCPSR people then tested the Arabic questionnaire and modified it in response to feedback received. The finalized version (appendix B) performed satisfactorily in a one-day pilot experiment in the Gaza Strip.

The essence of the questionnaire is to first build a roster of the respondent’s household members as of October 6, 2023, and then to account for each member’s post-October-7 fate, including whether they are alive, dead, or missing. A follow-up question asks respondents to divide reported deaths into violent and nonviolent ones.

PCPSR field teams have extensive experience in conducting public opinion surveys in the Gaza Strip. However, the GMS was their first mortality survey.

Accurate mortality estimation relies on partitioning the population into non-overlapping units for selection, ensuring that any death can enter the sample through only a single channel, i.e., the household to which the decedent belonged.

Accordingly, the pre-survey training for interview teams stressed the importance of respondents only being allowed to report household members (as of October 6, 2023) and not to report deaths of extended family members who did not live within the respondent’s household. This aspect of the training was, evidently, successful as the mean household size for the sample, 4.9, is rather lower than the mean of 5.5 reported by the PCBS (Palestinian Central Bureau of Statistics 2023).

Data collection began on December 30, 2024, and ended on January 5, 2025. At this stage, PCPSR deployed ten two-person teams, mostly females, of highly experienced data collectors. Four supervisors continuously monitored the fieldwork and attended more than a fifth of the interviews. A fieldwork coordinator ensured accurate deployment of the supervisors throughout the full workday.

Interviews were conducted using tablets or mobile phones that recorded GPS locations of each team during the data collection. A quality control team from PCPSR’s central office in Ramallah used these data to monitor team movements in the field. There was also continuous communication among PCPSR workers via WhatsApp.

All interviews were conducted face to face with adults aged 16 and above. Completed interviews were automatically uploaded to a secure, central server that can be accessed only by PCPSR researchers.

## 3. Data and Code

All data and code can be accessed at this URL: https://gitfront.io/r/mspagat/tZwP79d7Pntz/Gaza-Mortality-Survey/ along with supplementary tables, a sensitivity analysis and further information on sampling and fieldwork.

## Acknowledgements

We would like to thank Mr. Mr. Walid Ladadweh for supervising the sample selection and the deployment of the data collection team and the data collection team itself who must remain anonymous.

## Contributions

MS, DGS and JP conceived the project. DGS played a critical role in organizing the team and, along with HH, securing funding. MS, DGS, JP, KS and MR did the initial survey planning with all authors eventually contributing. All authors, but especially KS, monitored and provided feedback during data collection. JP, MS, KS and EB conducted analysis with all authors feeding back. KS and MR drafted the first description of the sampling and fieldwork. MS wrote the first draft of the main paper with EB playing a strong role in bringing it to completion. All authors contributed to, reviewed and approved all written material.

No authors of this paper have a competing interest.

## Footnotes

^1^On January 6, 2025 the GMoH put the death toll at 45,650 (12) however the GMoH’s list of violent deaths is a work in progress and the GMoH shared with the authors a recent version of the dataset with 49,090 deaths for the GMS coverage period.

^2^Updated GMoH data for the period covered by (7) place the comparable GMoH figure somewhat higher than it was when (7) was published. So, we have lowered their estimated percentage GMoH undercount from 41%, which was correct when (7) was published, to an updated 39.1%.

^3^Reassuringly, PCPSR population estimates across these three dwelling types remained reasonably stable over the three months preceding the survey.

