## Supplementary Information for "Violent and Nonviolent Death Tolls for the Gaza War: New Primary Evidence"

#### SUPPLEMENTARY MATERIALS

Supplementary Tables

Table S1: Mortality rates for the Gaza Strip October 6, 2023 to January 5, 2025

| Type | CDR | Standard error | Confidence Interval |
|---|---|---|---|
| Violent | 33.1 | 8.01 | (27.9, 38.4) |
| Nonviolent | 7.16 | 2.70 | (5.40, 8.93) |
| Total | 40.3 | 8.74 | (34.6, 46.0) |

Raked to age, gender, governorate, and household size

Based on household roster and births and IDB US Census Bureau population estimates

Deaths per 1,000 per year

Table S2: Estimates of violent and nonviolent mortality by governorate of residence on October 6, 2023

| Governorate | Estimate | Standard error | Confidence Interval |
|---|---|---|---|
| Total | | | |
| Northern Gaza | 16,000 | 3,230 | (9,630, 22,300) |
| Gaza city | 35,700 | 4,670 | (26,500, 44,800) |
| Deir al-Balah | 12,400 | 1,890 | (8,660, 16,100) |
| Khan Younis | 18,200 | 2,470 | (13,400, 23,000) |
| Rafah | 9,220 | 1,820 | (5,660, 12,800) |
| Total | 91,400 | 6,450 | (78,800, 104,000) |
| Violent deaths | | | |
| Northern Gaza | 13,200 | 2,850 | (7,620, 18,800) |
| Gaza city | 31,300 | 4,550 | (22,400, 40,200) |
| Deir al-Balah | 8,280 | 1,420 | (5,490, 11,100) |
| Khan Younis | 15,100 | 2,160 | (10,800, 19,300) |
| Rafah | 7,330 | 1,630 | (4,140, 10,500) |
| Total | 75,200 | 5,920 | (63,600, 86,800) |
| Nonviolent deaths | | | |
| Northern Gaza | 2,760 | 821 | (1,150, 4,370) |
| Gaza city | 4,370 | 975 | (2,460, 6,280) |
| Deir al-Balah | 4,100 | 1,170 | (1,800, 6,400) |
| Khan Younis | 3,130 | 890 | (1,390, 4,880) |
| Rafah | 1,890 | 704 | (509, 3,270) |
| Total | 16,300 | 2,040 | (12,300, 20,200) |

Raked to age, gender, governorate, and household size

Based on GMS Household roster.sav

Table S3: Gaza Strip population at risk 2023, broad age groups based on the International Data Base, U.S Census Bureau

| Age | Males | Male percent | Females | Female percent |
|---|---|---|---|---|
| 0-4 | 182,000 | 8.4 | 174,000 | 8.0 |
| 5-11 | 200,000 | 9.2 | 189,000 | 8.7 |
| 12-17 | 165,000 | 7.6 | 158,000 | 7.3 |
| 18-29 | 248,000 | 11.4 | 240,000 | 11.0 |
| 30-39 | 131,000 | 6.0 | 133,000 | 6.1 |
| 40-64 | 147,000 | 6.8 | 150,000 | 6.9 |
| 65+ | 29,900 | 1.4 | 28,700 | 1.3 |
| Total | 1,100,000 | 50.7 | 1,070,000 | 49.3 |

Percentages are based on the total population

Births during survey period added

Source: https://www.census.gov/ International Programs Main/ Data /International Database.

Downloaded 25.01.2025

Table S4: Gaza Strip Survey main status outcomes

| Status | Estimate | Standard error | Confidence Interval |
|---|---|---|---|
| Male | | | |
| Resident | 983,000 | 7,000 | (969,000, 997,000) |
| Left the Gaza Strip | 28,300 | 3,120 | (22,200, 34,400) |
| Elsewhere in the Gaza Strip | 15,100 | 2,260 | (10,700, 19,500) |
| Dead | 56,700 | 4,000 | (48,900, 64,500) |
| Missing | 11,100 | 1,980 | (7,190, 15,000) |
| Imprisoned | 9,420 | 1,670 | (6,150, 12,700) |
| Female | | | |
| Resident | 995,000 | 6,370 | (983,000, 1,010,000) |
| Left the Gaza Strip | 28,900 | 4,080 | (20,900, 36,900) |
| Elsewhere in the Gaza Strip | 11,900 | 2,160 | (7,690, 16,200) |
| Dead | 34,700 | 3,750 | (27,400, 42,100) |
| Missing | 1,140 | 559 | (42.0, 2,230) |
| Imprisoned | 157 | 160 | (-156, 470) |
| Total | | | |
| Resident | 1,980,000 | 11,500 | (1,960,000, 2,000,000) |
| Left the Gaza Strip | 57,200 | 6,020 | (45,400, 69,000) |
| Elsewhere in the Gaza Strip | 27,000 | 3,740 | (19,700, 34,400) |
| Dead | 91,400 | 6,450 | (78,800, 104,000) |
| Missing | 12,200 | 2,200 | (7,900, 16,500) |
| Imprisoned | 9,580 | 1,690 | (6,260, 12,900) |

Raked to age, gender, governorate, and household size

Based on Gaza Mortality Survey GMS Household roster.sav

Table S5: Estimates of missing persons by broad age and gender groups

| | Estimate | Standard error | Confidence Interval |
|---|---|---|---|
| Missing persons | | | |
| Children (<18) | 3,610 | 1,040 | (1,570, 5,650) |
| Women (18-64) | 131 | 134 | (-133, 394) |
| Elderly (65+) | 0 | 0 | (0, 0) |
| Men (18-64) | 8,470 | 1,650 | (5,240, 11,700) |
| Total missing persons | 12,200 | 2,200 | (7,900, 16,500) |
| Women, elderly, and children | 30.6 | 6.40 | (18.1, 43.2) |

Raked to age, gender, governorate, and household size
Based on GMS Household roster.sav

Table S5a: Gaza Strip Survey Non violent deaths by child/adult status and gender

| Child/adult | Deaths | Standard error | Confidence interval |
|---|---|---|---|
| Men | | | |
| Adults | 5,210 | 1,180 | (2,900, 7,510) |
| Children<18 | 1,680 | 741 | (229, 3,130) |
| Women | | | |
| Adults | 8,250 | 1,350 | (5,590, 10,900) |
| Children<18 | 1,120 | 579 | (-17.3, 2,250) |
| Total | | | |
| Adults | 13,500 | 1,760 | (10,000, 16,900) |
| Children<18 | 2,800 | 925 | (986, 4,610) |

Raked to age, gender, governorate, and household size
Based on Gaza Mortality Survey GMS Household roster.sav

#### Sensitivity Analysis

Table S6: Sensitivity of estimates to model assumptions

| | Main model | | Team 9 excluded | | Census 2017 household size adjustment | | Full rake, no weight trim | | PCBS population, full rake | | No household size adjustment | | Unadjusted equal probability | |
|---|---|---|---|---|---|---|---|---|---|---|---|---|---|---|
| Categories | Estimate | SE[1] | Estimate | SE | Estimate | SE | Estimate | SE | Estimate | SE | Estimate | SE | Estimate | SE |
| Violent deaths | | | | | | | | | | | | | | |
| Children (<18) | 22,800 | 3,090 | 19,200 | 2,810 | 22,400 | 3,040 | 22,800 | 3,080 | 24,100 | 3,260 | 25,800 | 3,600 | 21,100 | 2,850 |
| Women (18-64) | 16,600 | 2,220 | 13,200 | 2,090 | 16,000 | 2,140 | 16,100 | 2,170 | 17,100 | 2,300 | 17,700 | 2,300 | 21,400 | 2,810 |
| Elderly (65+) | 2,870 | 943 | 3,060 | 1,110 | 3,440 | 1,200 | 3,350 | 1,170 | 3,550 | 1,240 | 2,800 | 909 | 1,940 | 637 |
| Men (18-64) | 32,900 | 2,700 | 28,700 | 2,640 | 32,000 | 2,640 | 32,100 | 2,640 | 34,000 | 2,800 | 33,600 | 2,740 | 40,800 | 3,330 |
| Total violent deaths | 75,200 | 5,920 | 64,100 | 5,410 | 73,800 | 5,780 | 74,400 | 5,860 | 78,700 | 6,200 | 79,900 | 6,640 | 85,200 | 6,710 |
| Violent deaths % | | | | | | | | | | | | | | |
| Women, elderly, and children | 56.2 | 2.94 | 55.2 | 3.39 | 56.6 | 2.94 | 56.9 | 2.94 | 56.9 | 2.94 | 57.9 | 2.88 | 52.2 | 2.89 |
| Nonviolent deaths | | | | | | | | | | | | | | |
| Total nonviolent deaths | 16,300 | 2,040 | 17,500 | 2,230 | 17,200 | 2,240 | 17,300 | 2,230 | 18,300 | 2,360 | 16,200 | 2,020 | 14,200 | 1,840 |
| Excess nonviolent deaths | 8,540 | 2,040 | 9,790 | 2,230 | 9,510 | 2,240 | 9,600 | 2,230 | 10,600 | 2,360 | 8,520 | 2,020 | 6,520 | 1,840 |
| Missing persons | | | | | | | | | | | | | | |
| Children (<18) | 3,610 | 1,040 | 3,300 | 1,010 | 3,510 | 1,020 | 3,540 | 1,020 | 3,750 | 1,080 | 3,960 | 1,120 | 3,020 | 828 |
| Women (18-64) | 131 | 134 | 136 | 139 | 125 | 129 | 126 | 129 | 133 | 137 | 194 | 196 | 216 | 216 |
| Elderly (65+) | 0 | 0 | 0 | 0 | 0 | 0 | 0 | 0 | 0 | 0 | 0 | 0 | 0 | 0 |
| Men (18-64) | 8,470 | 1,650 | 8,360 | 1,710 | 8,100 | 1,560 | 8,230 | 1,600 | 8,720 | 1,700 | 8,720 | 1,790 | 10,100 | 1,980 |
| Total missing persons | 12,200 | 2,200 | 11,800 | 2,270 | 11,700 | 2,090 | 11,900 | 2,140 | 12,600 | 2,270 | 12,900 | 2,520 | 13,400 | 2,430 |
| Missing persons % | | | | | | | | | | | | | | |
| Women, elderly, and children | 30.6 | 6.40 | 29.1 | 6.36 | 31.0 | 6.55 | 30.8 | 6.43 | 30.8 | 6.43 | 32.3 | 6.07 | 24.2 | 5.17 |

[1]SE: Standard error

Based on GMS Household roster.sav

Table S6A. Scheme showing the cases for the models in table S6

| Model | USCB population projection | PCBS population projection | MICS Household size | Census 2017 Household size | Age distribution | Gender | Governorate of origin | Trim |
|---|---|---|---|---|---|---|---|---|
| Main | ■ | | ■ | | ■ | ■ | ■ | ■ |
| Team 9 excluded | ■ | | ■ | | ■ | ■ | ■ | |
| Census 2017 adjustment for household size | ■ | | | ■ | ■ | ■ | ■ | |
| Full rake, no trim | ■ | | ■ | | ■ | ■ | ■ | |
| PCBS Population, full rake | | ■ | ■ | | ■ | ■ | ■ | |
| No household size adjustment | ■ | | | | ■ | ■ | ■ | |
| Unadjusted equal probability | ■ | | | | | | | |

Table S6 gives results from six new estimations, each one departing from some of the assumptions made in the main model. Table S6A displays the differences between the cases. Only one model uses the estimate for the total population of the Gaza Strip made by the Palestinian Central Bureau of Statistics (PCBS). All other models use the US Census Bureau (UCB) total population estimates. Four models use a Multiple Indicator Cluster Survey (MICS) of the Gaza Strip to weight for household size while one weights for household size using UCB data and two models do not weight for household size at all. All but one model weight for gender, age and governorate of origin using UCB data while one model presents unweighted estimate. None of the six new models trims relatively extreme weights to back to more moderate values (1).

Only two models produce estimates that differ substantially for those of the main model and the other four models, "Team 9 excluded" and "Unadjusted equal probability", so we consider each one in greater depth below.

The survey used ten interview teams and one, known as "gaza9", recorded 100 out of the 393 violent deaths in the sample. So, we ran a model that excludes the gaza9 data. Compared to the main estimates this scenario (a) decreases the central estimate for violent deaths by more than 10,000 deaths, from 75,200 to 64,100; (b) slightly changes the central estimate for the percentage of women, children, and elderly among the violent deaths from 56.2% to 55.2%; (c) increases nonviolent excess deaths from 8,540 to 9,790. Thus, exclusion of the gaza9 data does not affect our main qualitative conclusions regarding the fact of MoH undercount, the percentage of women, children, and elderly or excess nonviolent deaths.

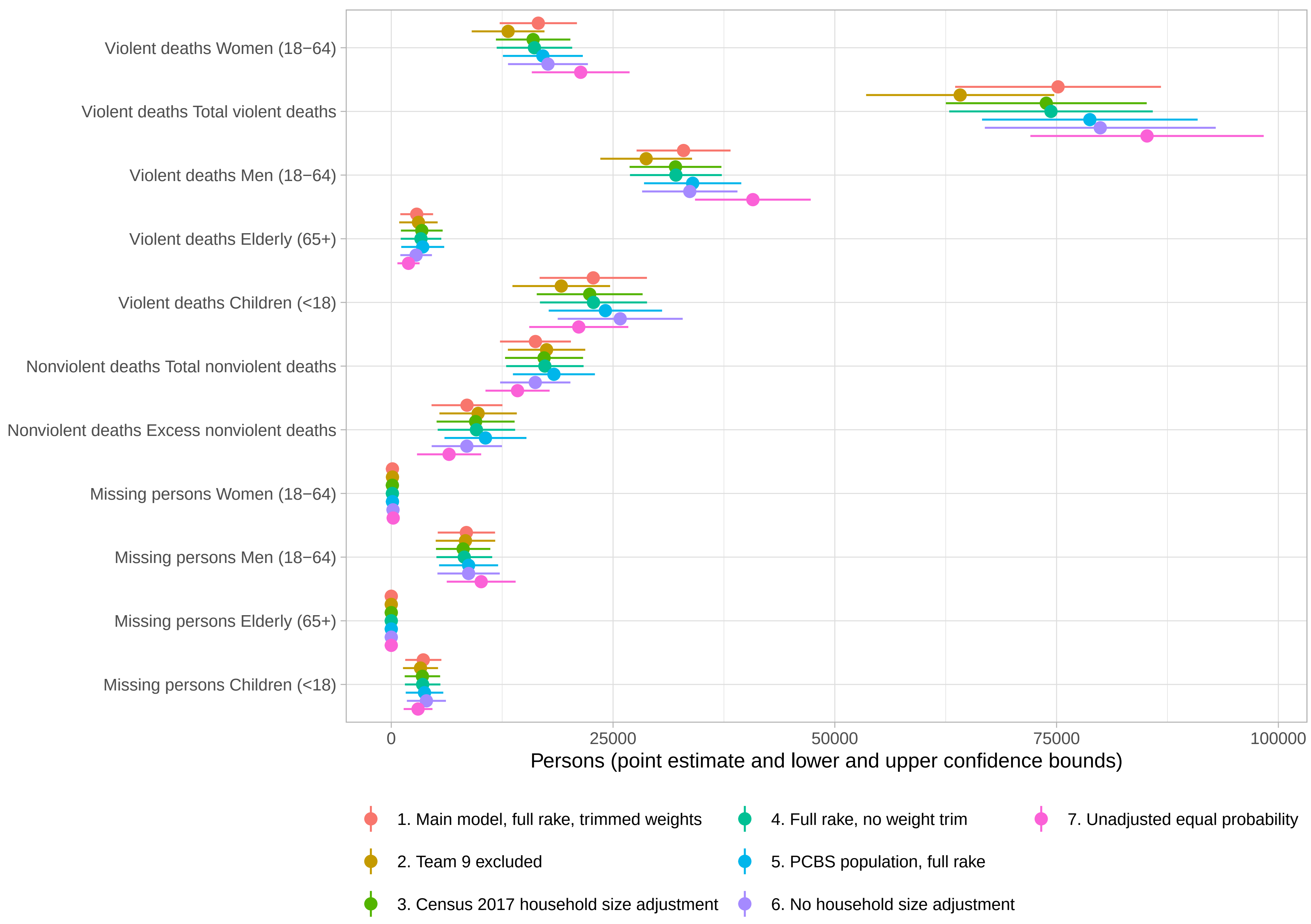
Violent deaths Women (18−64)
Violent deaths Total violent deaths
Violent deaths Men (18−64)
Violent deaths Elderly (65+)
Violent deaths Children (<18)
Nonviolent deaths Total nonviolent deaths
Nonviolent deaths Excess nonviolent deaths
Missing persons Women (18−64)
Missing persons Men (18−64)
Missing persons Elderly (65+)
Missing persons Children (<18)
0
25000
50000
75000
100000
Persons (point estimate and lower and upper confidence bounds)
1. Main model, full rake, trimmed weights
2. Team 9 excluded
3. Census 2017 household size adjustment
4. Full rake, no weight trim
5. PCBS population, full rake
6. No household size adjustment
7. Unadjusted equal probability

Yet, omission of the data collected by the gaza9 team does substantially lower our estimate for the size of the MoH undercount. So, we investigated further and found that three PSUs covered by this team were in shelters that give special preference to families that have lost members during the fighting, especially to those who have lost one or both parents. An additional five gaza9 PSUs were in or near Bani Suheila, Mawasi Rafah or the northern border of Khan Yunis, all areas that have experienced heavy attacks. Thus, it appears that the high numbers recorded by the gaza9 team come mainly from sampling variation. Moreover, comparison of interviews with and without supervisors for this team reveal no discrepancies. Thus, we are confident in the quality of these interviews and just note their impact here.

The only other model that produced estimates that differ substantially from the main model is the one based on just the raw data without weighting on anything, i.e., the so-called “unadjusted equal probability model”. This model estimates (a) 7,000 more violent deaths than the main model (85,200 vs. 75,200); (b) 4 percentage points fewer women, children and elderly (52.2% vs. 56.2%); (c) 2,010 fewer nonviolent deaths (14,200 vs. 16,300). These effects come largely from overrepresentation of men and small households in the sample compared to the population from which it is drawn. Thus, it is clear that the weighting scheme, as a whole, is important but we are not tempted to adopt the unadjusted equal probability model as the main model because transforming the sample to bring it into line with population characteristics should bring our estimates closer to the truth than they can be when based on just the raw numbers.

The final point from this sensitivity analysis worth noting is that the final stage of our raking (1) procedures, i.e., the one that trims the most extreme weights back to more moderate values, has no substantial impact on the estimates and could easily be dropped. In particular, the model “Full rake, no weight trim” produces estimates that hardly differ from those of the main model. However, weight trimming is standard procedure so we have left it in the main model.

The sensitivity plot on the next page displays much of the information in table S6.

#### Note on Raking

Table S7 (next page) first shows how raking transforms the sample to better match population characteristics. Its second column (“Sample Percentages”) gives demographic, locational and household-size characteristics of the sample. The next column (“Population Percentages”) shows the same characteristics but for the population of the Gaza Strip. The third column is the transformed (after raking) sample percentages (“Raked Sample”). The final column shows differences between population and raked values.

Unraked sample proportions are close to population proportions on demographics and governorate of origin. However, the two differ markedly on the distribution of household sizes. Raking brings everything into close alignment except for the percentage of households with ten or more members for which there is a difference of slightly more than 2 percentage points even after raking.

In the raw, i.e., unraked, data 3.1% of people in households with 10 or more pre-October-7 members died violently whereas for the other households the violent death rate was 4.0%. So further upweighting the 10+ would decrease our violent death estimate, but only slightly. Thus, this outlier of the raking process is not influential.

Table S7: Comparison of Population Values with Sample and Raked Sample Values

By Demographics, Governorate and Household Size

| Categories | Sample Percentages | Population Percentages | Raked Sample Percentages | Population - Raked Sample |
|---|---|---|---|---|
| Ages/Sexes | | | | |
| Male 0-4 | 6.96 | 8.67 | 8.42 | 0.257 |
| Female 0-4 | 6.25 | 8.29 | 7.53 | 0.759 |
| Male 5-14 | 7.94 | 9.54 | 9.20 | 0.341 |
| Female 5-14 | 7.09 | 9.01 | 8.39 | 0.615 |
| Male 15-17 | 5.80 | 7.88 | 6.94 | 0.943 |
| Female 15-17 | 6.06 | 7.52 | 7.65 | −0.124 |
| Male 18-29 | 12.5 | 11.8 | 11.1 | 0.771 |
| Female 18-29 | 14.0 | 11.4 | 11.9 | −0.473 |
| Male 30-39 | 7.57 | 6.22 | 6.32 | −0.102 |
| Female 30-39 | 6.85 | 6.34 | 6.04 | 0.298 |
| Male 40-64 | 9.09 | 7.00 | 7.48 | −0.472 |
| Female 40-64 | 7.95 | 7.15 | 6.57 | 0.574 |
| Male 65+ | 1.00 | 1.43 | 1.31 | 0.115 |
| Female 65+ | 0.863 | 1.37 | 1.18 | 0.190 |
| Governorates | | | | |
| North Gaza | 17.8 | 20.0 | 19.5 | 0.453 |
| Gaza City | 36.1 | 33.6 | 33.9 | −0.284 |
| Deir al Balah | 12.8 | 14.3 | 14.2 | 0.141 |
| Khan Younis | 20.8 | 19.7 | 19.8 | −0.151 |
| Rafah | 12.5 | 12.4 | 12.5 | −0.160 |
| Household Sizes | | | | |
| 1 | 2.05 | 0.577 | 0.572 | 0.00562 |
| 2 | 9.20 | 2.63 | 2.72 | −0.0873 |
| 3 | 14.8 | 5.61 | 5.82 | −0.210 |
| 4 | 17.0 | 9.66 | 9.98 | −0.323 |
| 5 | 18.6 | 12.9 | 13.3 | −0.440 |
| 6 | 15.0 | 16.9 | 17.3 | −0.429 |
| 7 | 11.2 | 17.3 | 17.6 | −0.372 |
| 8 | 5.45 | 11.8 | 11.9 | −0.104 |
| 9 | 3.95 | 9.53 | 9.64 | −0.115 |
| 10+ | 2.80 | 13.1 | 11.1 | 2.07 |

Notes on the Calculations

The computer code accompanying the paper spells out each step in our calculations. We recognize, however, that some else's computer code is hard to read so here we provide a short document in Q&A form that should clarify the essential steps.

**How do you make central estimates?**

Every individual in the sample is assigned a weight which gives, in essence, the number of people in the Gaza Strip that this individual represents.

Once these weights are in place it is simple to make central estimates. For example, the central estimate for violent deaths is the sum of the weights for all individuals in the sample recorded as dying violently.

**How are the weights calculated?**

The weights have two distinct components. First, is a multiplier to scale the sample size up to the full population of the Gaza Strip. There are 9,729 individuals in the sample who were alive on October 6, 2024. In addition, we recorded 357 births during the war. The UCB estimate for the population of the Gaza Strip is just over 2,089,389. So, on average, each individual in the sample represents somewhat more than 200 Gazans.

The second component of the weights raise or lower the scaling factor for an individual depending on whether the characteristics of that individual are, respectively, less or more common than these characteristics are in the full population of the Gaza Strip. So, if, for example, the proportion of women in the sample aged 40-64 is lower than the proportion of this group in the population then the weights on individuals from this group are increased above the scaling factor. Weights are adjusted in this way for gender, age, household size and governorate to bring weighted sample proportions broadly into line with population proportions. Finally, extreme weights are trimmed back to twice the median weight for the whole sample, although the sensitivity analysis (above) shows that trimming did not have a substantial impact on the estimates.

In the first calculations we made on the data we went through characteristic by characteristic and assigned a weight for each one to match sample and population proportions. We then multiplied these weights together to get final weights. So, for example, the final weight on, e.g., a 14-year-old female in a household of size 3 in Rafah governorate was the female 12-17 weight times the household-size-3 weight times the Rafah weight.

This procedure has the virtue of being easy to understand but the drawback is that when you weight for one characteristic you can throw the weights for other characteristics out of whack. So, we used *raking*, which is an iterative procedure that brings all included sample proportions roughly into line with population proportions.

Table S7 shows that this raking procedure was a success, but it is comforting to know that the raked results came out very close to the original ones where we weighted independently on each characteristic.

**What is the purpose of weighting on sex, age, household size and governorate?**

We weight to make our sample look, as much as possible, like a microcosm of the population it is drawn from. A random sample should, theoretically, come reasonably close to accomplishing this goal. However, in a sample of 2,000 there are bound to be some important deviations even if it is drawn under pristine conditions. So estimates can be improved by reshaping the sample so that the weight of people with characteristics that are under(over) represented in the sample are scaled up(down). Our sample was certainly not drawn under pristine conditions, so weighting was an important tool to make our sample look like a microcosm of the population.

**How are confidence intervals calculated?**

As noted above, the central estimates for death tolls are based on ratios, e.g., the number in the sample reported as dead divided by the total number of people in the sample. Nonlinear functions complicate variance estimation, the key step in calculating confidence intervals, so we (really the Survey package in R) use Taylor series to make linear approximations for our ratios. These linear approximations are not used for the central estimates themselves but, rather, for variance estimation and confidence intervals. The basic idea is that an estimated ratio is approximately equal to the true population ratio plus deviations in the numerator multiplied by the first derivative of the ratio with respect to changes in the numerator plus deviations in the denominator multiplied by the first derivative of the ratio with respect to changes in the denominator. For details see chapters 3 and 5 of:

Lumley, T. (2010). Complex Surveys: A Guide to Analysis Using R. Wiley.

It is worth noting that we also calculated confidence intervals using bootstrapping and got similar results. We haven't included these results and code because there is already quite a lot of supplementary material but we could add it if the editors want us to do so.

**How are excess deaths calculated?**

The US Census Bureau made projections for the number of deaths that would occur in the Gaza Strip during the period covered by the survey. These projections were made before the war and assumed that there would be no war. We simply subtracted US Census Bureau-based projected deaths from our central estimate for nonviolent deaths to get our central estimate for excess non-violent deaths. The upper (lower) limit of the 95% confidence interval for nonviolent excess deaths is the upper (lower) limit of the 95% confidence interval for nonviolent deaths minus US Census Bureau-based projected deaths.

#### Note on GMoH Data Quality

Since October 7, 2024, the Gaza Ministry of Health (GMoH) has tried to make an individualized and public record for each Gazan killed violently in the war. The fruit of this casualty recording project has been a series of detailed lists of Gazans documented to have been killed in the war. The most recent list, covering the period from the beginning of the war through May 11, 2025, contains 52,958 entries.

The credibility of the GMoH lists is underpinned, first and foremost, by their exceptional transparency. They openly list the name, ID number, sex and age of each victim. Israel maintains a population registry for the Gaza Strip that enables it to verify the authenticity of each listed person but has not challenged these open accounts. And many of the people listed by the GMoH have been verified as dead through social media monitoring, notably in this study.

Many Gazans have been killed very rapidly in the war. Moreover, the GMoH has limited resources to record all these deaths, and their capacity constraints are exacerbated by attacks on hospitals. Thus, it is impossible for the GMoH to record every single death. These challenging conditions have led to many flaws, such as incomplete entries or entries with invalid ID numbers, in the GMoH's published records, particularly during the first half of 2024. However, these flaws have been cleaned up over time and are no longer a significant factor affecting the quality of the records.

The essays collected here plus this recent survey track the trajectory over time in GMoH data quality and provide a basis for understanding both the high value of the GMoH records as well as the remaining puzzles to be resolved. For present purposes, the main conclusions to draw from research into GMoH data quality is that these records are reliable but not comprehensive. Thus, we should regard the total number of fatalities recorded by the GMoH as a reliable, indeed a conservative, lower bound for the total number of people killed in the war.

#### An In-depth View of the Sampling, Fieldwork and Data Cleaning

PCPSR's sample for the mortality rate survey was designed to be representative of Gazan residents, taking into account what is feasible with the ongoing conflict. The sample design includes stratification at the first stage and two hundred Primary Sampling Units (PSUs) representing all governorates of the Gaza Strip were selected based on area probability sampling, from a frame of "enumeration areas" and shelters, or displacement centers. These enumeration areas and shelters were located in two governorates: Khan Younis and Central Gaza Strip (or Deir al Balah). Three Gazan governorates were inaccessible due to active combat or Israeli standing evacuation orders: Northern Gaza, Gaza City, and Rafah. Any Gazans currently living in these areas were excluded from the sample. However, due to Israeli military action and evacuation orders, the vast majority of Gazans have been displaced from these areas yielding a limited proportion of the population in each at the time of the survey. In August 2024, international organizations estimated that 90% of Gazans have been forced to relocate from their homes. Since then, many more people in the northern Gaza Strip were forced to relocate to the south, which includes the governorates where the survey was fielded.

Available population estimates indicate that the vast majority of the displaced persons from these three inaccessible governorates were sheltering in built up or tent shelters in the two accessible governorates. Accordingly, by interviewing those who were displaced, it is possible to develop estimates of violent and excess mortality for these areas of the Gaza Strip. Additionally, many of the displaced persons from the two accessible governorates were able to return to their own homes in these governorates after the termination of active combat and the withdrawal of the Israeli army from these areas, meaning few remained in built-up shelters or tent shelters. These formerly displaced persons were interviewed in their own homes in the "enumeration areas."

To determine the actual distribution of the population in each shelter, PCPSR works closely with three actors: 1) governmental sources, 2) the UN and UNRWA, and 3) relevant international and non-governmental actors. Satellite images provide additional information to confirm locations. PCPSR's own fieldwork force in the Gaza Strip keeps a record of all the shelters and their population which they update through updated satellite images and contacts with the local municipal authorities by our data collectors. PCPSR data collectors visit all PSUs and confirm the most updated locations and numbers by talking directly to those managing the shelters. Records have been updated every three months since the start of the Israeli military campaign to reflect the changed shelter landscape within the Gaza Strip.

Within each PSU, ten households were randomly selected for a total sample of 2,000 households. The total number of household members in these households on October 6, 2023, stood at 9,729. All interviews were conducted face-to-face with adults aged 16 and above, regardless of the adult age or the gender. In total, 58 eligible families, of which 42 were in shelters and 16 in enumeration areas, refused to be interviewed. This yields a response rate for eligible respondents of over 97%.

Table S8. Households and household members by governorate: sample versus PCBS 2023 data

| Governorates | PCBS 2023 Population | % of population | number of households | % of households | number of household members | % of household members |
|---|---|---|---|---|---|---|
| Northern Gaza | 444,412 | 20.0 | 360 | 18.0 | 1,735 | 17.8 |
| Gaza City | 749,100 | 33.6 | 716 | 35.8 | 3,525 | 36.2 |
| Khan Younis | 438,557 | 19.7 | 420 | 21.0 | 2,034 | 20.9 |
| Deir al-Balah | 319,208 | 14.3 | 279 | 14.0 | 1,216 | 12.5 |
| Rafah | 275,267 | 12.4 | 225 | 11.3 | 1,219 | 12.5 |
| Total Gaza Strip | 2,226,544 | 100.0 | 2,000 | 100.0 | 9,729 | 100.0 |

Data: Official data (Palestinian Central Bureau of Statistics 2021) and GMS data.

**Strata and PSUs**:

The sample was allocated to three primary strata based on available population estimates from the Palestinian Central Bureau of Statistics (PCBS) and estimates informed by the UN / UNRWA and relevant international and non-governmental actors. These three strata are: enumeration areas, built up shelters, and tent shelters. Governorates, based on residency on October 6, 2023, served as a substratum for each of the three primary strata. Within each stratum, PSUs were allocated in proportion to the estimated population within each. In each PSU, 10 households were randomly selected. Within each household, the respondent was not selected at random. Instead, interviews were conducted with a willing adult (ages 16+) regardless of age or gender of the respondent. This approach was implemented to minimize non-response rates as any competent adult within the household is expected to be able to enumerate the number and current status of household members living with them on October 6, 2023.

1) In the **enumeration areas**, representing the first stratum, interviews were conducted in the two accessible governorates, Khan Younis and Deir al Balah, in populated areas specified by the PCBS as a representative sample of these governorates. Only permanent residents of these two governorates, based on October 6, 2023 residency location, were eligible for selection within this stratum. The sample for these areas, along with its associated detailed unit-by-unit maps, was originally purchased by PCPSR from PCBS's 2017 census, the latest census conducted in the Palestinian territories. Each of these 70 selected areas represent a PSU. The total number of households where interviews were conducted stands at 699 for a total of 3250 household members.[1] The households were selected in enumeration areas by dividing the total number of units in each enumeration area over the number of interviews. The resulting number is the interval used to determine the location of the next home to be interviewed once the first interview was conducted.

2) In the **built-up shelters**, mostly schools, universities, clubs and other standing areas that have not been destroyed during the war, a regular random sample was drawn from the lists of blocks representing all the shelters in each governorate of Khan Younis and Deir al Balah. Each shelter was divided into blocks of roughly equal size and 41 PSUs were allocated to this stratum. These 41 PSUs were allocated proportionally across all the shelters, meaning multiple blocks were allocated to shelters with a particularly large number of internally displaced persons. This process resulted in the selection of 22 shelters in Khan Younis and 17 shelters in Deir al Balah for a total of 39 shelters.

Because the shelters contain internally displaced residents, a substratum based on governorate of residence on October 6, 2023 was applied to ensure greater representation. Only those who were residents of the three inaccessible governorates – Northern Gaza, Gaza City, and Rafah – were eligible for selection in built-up shelters. Interviews were allocated to each substratum based on the relative proportion of each of these three governorates on October 6, 2023. This approach

[1] In rare cases, interviews were dropped for eligibility or quality control issues. In such cases, additional interviews were sometimes collected in different areas to compensate when the team had already left the original enumeration area. As a result, the number of interviews in PSU is not always exactly 10.

was designed to ensure a fair representation of the population from the original governorates and localities (such as Northern Gaza) from which they came.

The process for interviews was as follows. The field team working in built-up shelters was informed of the allocation of each PSU to residents from a specified governorate. If a contacted household was from that governorate, then the interview process was initiated. If the household was not living in the selected governorate on October 6, 2023, then the household was deemed ineligible and the standard skip pattern applied for selecting a new household. Once the PSU reached ten eligible households that agreed to participate, the PSU was considered complete. This process was then repeated for the next PSU.

Additionally, hundreds of small gatherings, dozens of families each, were identified around these shelters and were included in our sampling frame by including them in the PSU of the nearest shelter to each such locality. The interviews were conducted inside 408 households in these built-up shelters for a total of 2,076 household members. The calculation of the interval between households in all shelters is the same as that of the enumeration areas.

3) In the **tent shelters** in the governorates of Khan Younis and Deir al Balah, PCPSR fieldwork teams, deployed on the ground since the war started, identified 65 major tent shelters in Khan Younis and 51 in Deir al Balah for a total of 116 shelters. A total of 89 PSUs were allocated to this stratum. Using the same methodology as in the case of the built-up shelters, these areas were divided into blocks of similar sizes and a regular random sample of 89 blocks were drawn. The same process of sub-stratification by the governorate of residence on October 6, 2023, was employed as in the case of built-up shelters with only those from the three inaccessible governorates eligible for inclusion. Interviews with 893 households were conducted inside the tents for a total of 4,403 household members.

Table S9: PSU type by household units

| | | Frequency | Percent | Valid Percent | Cumulative Percent |
|---|---|---|---|---|---|
| Valid | Built up shelter | 408 | 21.4 | 20.4 | 21.4 |
| | Tent shelters | 893 | 43.7 | 44.7 | 65.1 |
| | enumeration area | 699 | 35.0 | 35.0 | 100.0 |
| | Total | 2000 | 100.0 | 100.0 | |

Table S10: PSU type by household members

| | | Frequency | Percent | Valid Percent | Cumulative Percent |
|---|---|---|---|---|---|
| Valid | Built up shelter | 2076 | 20.9 | 21.3 | 20.9 |
| | Tent shelters | 4403 | 45.8 | 45.3 | 66.6 |
| | enumeration area | 3250 | 33.4 | 33.4 | 100.0 |
| | Total | 9729 | 100.0 | 100.0 | |

Table S11: Distribution of PSUs by governorates and type

| | | PSU type by households | | | | | | |
|---|---|---|---|---|---|---|---|---|
| | | Built up shelter | | Tent shelters | | enumeration area | | Total |
| Governorates | Northern Gaza | 230 | 63.9% | 130 | 36.1% | | | 360 |
| | Gaza City | 158 | 22.2% | 558 | 77.8% | | | 716 |
| | Khan Younis | | | | | 420 | | 420 |
| | Central Gaza Strip (Deir al Balah) | | | | | 279 | | 279 |
| | Rafah | 20 | 8.9% | 205 | 91.1% | | | 225 |
| | Total Gaza Strip | 408 | 20.4% | 893 | 44.7% | 699 | 35.0% | 2000 |

**Data collection and quality control:**

Data collection started on 30 December 2024 and ended on 5 January 2025. The demographics of the respondents as of October 6, 2023 were as follows: 61% male, 39% female, 12% aged 16-24, 32% 25-34, 25% 35-44, 17% 45-54 and 14% 55+.

Ten teams of highly experienced data collectors, mostly females, were deployed each day. Each interview was conducted by two data collectors. Four supervisors were deployed to monitor fieldwork every day to ensure accurate deployment of the ten data collection teams. At least one third of the interviews were conducted in the presence of a supervisor. One fieldwork coordinator was deployed to ensure the accurate deployment of all supervisors throughout the full workday.

Interviews were conducted by using tablets or mobile phones that recorded the GPS location of teams during the data collection. PCPSR staff and the quality control team based in PCPSR's central office monitored live the movement of the data collectors by plotting the GPS locations on a map of the Gaza Strip. When each interview was completed, it was automatically sent directly to our secure, central server where only PCPSR researchers have access to it.

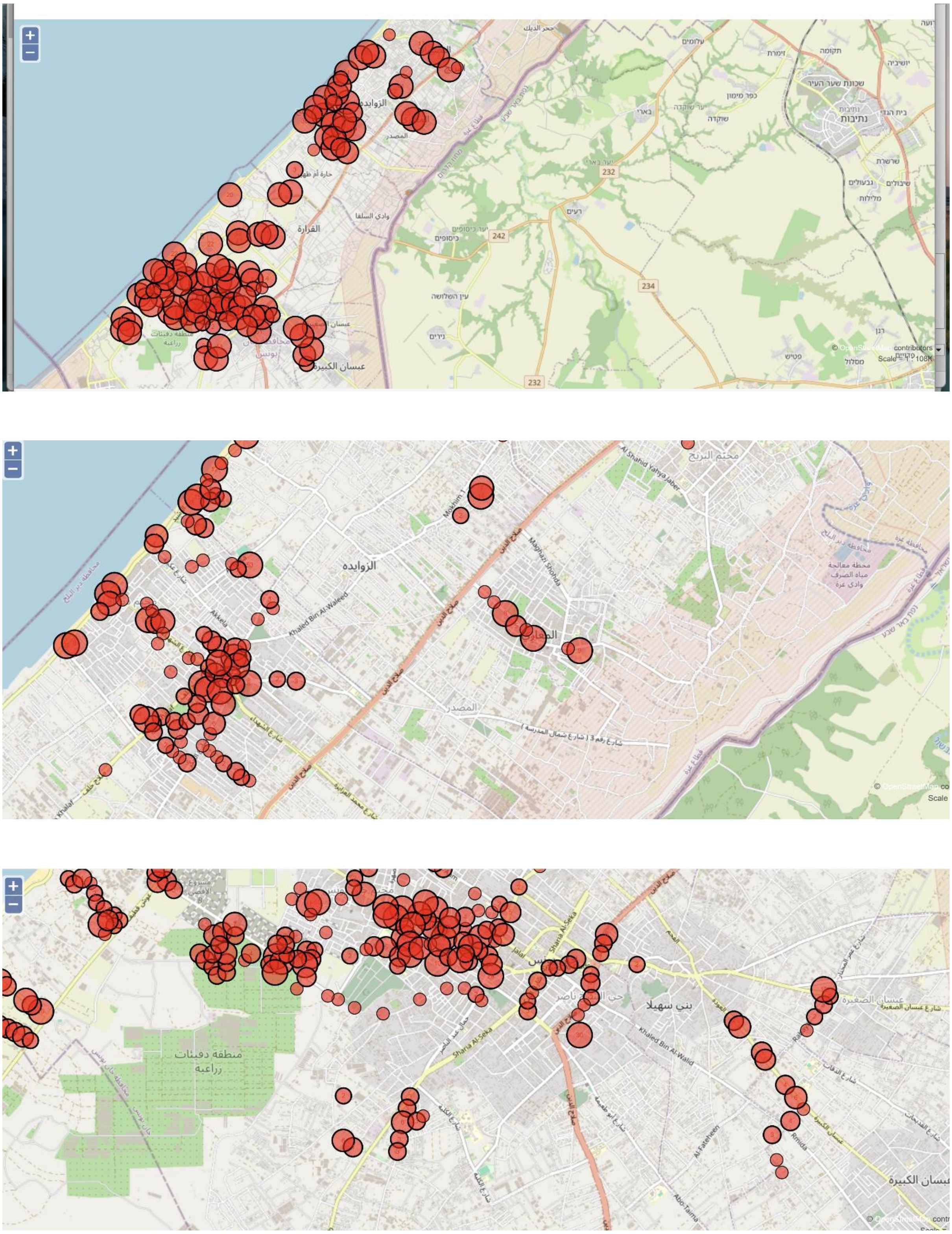

Figure S2. Locations of the interview teams during the survey.

All teams, monitors, coordinator, and central research teams were connected via two WhatsApp groups that ensured full simultaneous communication with the teams to resolve any emerging problems.

To ensure the safety of our data collectors in the Gaza Strip, interviews were conducted with residents in areas that did not witness active combat and were not designated as an evacuation area during the period of fieldwork.

##### Data Cleaning

Here are the steps in the data cleaning process:

1. Matching the number of PSUs, interviews, localities with the required sample size and locations
2. Matching question texts and answer options with questionnaire questions
3. Review the duration of the interview in terms of start and end time
4. Matching the team numbers with the counting areas/localities specified in the deployment plan
5. Matching the geographical locations, or GPS, with the geographic deployment plan
6. Matching the names of residential communities and governorates with those in the sample specification
7. Ensuring no duplication of data files during the transfer process from tablets to server
8. Review and address any outliers in answers to all questions
9. Review logical errors in answers to various questions
10. Review errors in related questions, such as skips, or follow ups, if any
11. Check the conformity of specific demographic questions, such as age or family size, with official data from PCBS

### Survey on mortality in the Gaza Strip after October 6, 2023

For the researcher: Ask the respondent about the governorate in which their family lived on 6th of October (in shelters, make sure the respondent's governorate is included in your team's sample. If it is not, move on to the next family until you find a family from the governorate included in your team's sample):

d04 Governorate Name:

1. Jabalia, North Gaza

2. Gaza City

3. Khan Yunis

4. Deir al-Balah

5. Rafah

#### Questionnaire

##### Section 1: Identification

| | | | |
|---|---|---|---|
| ID00 | **Team Number** | | |
| ID01 | **Household number** | To be filled in by CAPI | \|__\|__\|__\|__\| |
| ID02 | **Location number** | To be filled in by CAPI | \|__\|__\|__\| |
| ID03 | **Stratum** | To be filled in by CAPI | \|__\|__\| |
| ID04 | **Date of interview (yyyy/mm/dd)** | To be filled in by CAPI | ______/__/__ |
| ID05 | **Time of interview start** | To be filled in by CAPI | __:__ |
| ID06 | **GPS coordinates of the interview location** | To be filled in by CAPI | |
| ID07 | **Interviewer** | To be filled in by CAPI | \|__\|__\| |
| ID08 | **Supervisor name** | | |

##### Section 2: Informed consent

***Thank you for taking the time to speak to me. The reason for contacting you is that we (organization) are conducting a survey of the number of deaths that occurred after the current hostilities. We carry out the study by asking a sample of persons about the members of their households and any deaths that have occurred after October 6, 2023. You have been selected as a respondent by chance. While we will ask about your name and the names of your household***

***members during the interview, we will not retain the names after we have completed the interview. We will also not record any information that can be used to identify you after the interview, either by us or by anyone else.***

***You may ask me to stop the interview any time if you wish.***

| | | | |
|---|---|---|---|
| CO01 | **Are you comfortable with us starting the interview? Would you like to refrain from answering? or do you have any questions before we start?** | 1: Can start<br><br>2: Questions, start after<br><br>3: Questions, refusal<br><br>4: Refusal | *à End interview (ID08)*<br><br>*à End interview (ID08)* |

#### Section 3: Household location on October 6th, 2023 and composition

| | | |
|---|---|---|
| HR00 | **In what governorate of the Gaza Strip did your household live on October 6, 2023?** | 1: North Gaza<br><br>2: Gaza City, north of Israeli road (Netzarim corridor)<br><br>3: Khan Younis<br><br>4: Central Gaza: south of Israeli road (Netzarim corridor) Deir el Balah, Nusairat, al Buraij, and al Maghazi<br><br>5: Rafah |

***Please provide information on each household member you lived with on October 6, 2023. I will also ask about each person's present situation. Start with yourself.***

| HR01: **Serial number of household member** | HR02: **Name of household member.** (The name will only be used for reference during the interview and will | HR03: **How old was …. on October 6, 2023?** | HR04: **Gender**<br>1: Male<br>2: female | HR05: **Is …. still living in the household, or has he/she moved, died, or is missing**<br>1 . Still resident à Next person | HR06: **Cause of death**<br>1. Disease, old age, cancer, pre-existing conditions etc<br>2. Disease, old age, cancer, pre-existing conditions etc not receiving medical aid |
|---|---|---|---|---|---|

| | | | | | |
|---|---|---|---|---|---|
| | not be included in the data file) | One or two digits,<br>95: 95+<br>98: Don't know<br>99: Refused | | 2. Left Gaza à Next person<br>3. Moved within Gaza à Next person<br>4. Dead<br>5. Missing<br>6. Imprisoned<br>à Next person | 3. Accident<br>4. Killed in/because of fighting<br>5. Do not know |
| 1 | | | | | 1 à *Next person* |
| 2 | | | | | |
| 3 | | | | | |
| 4 | | | | | |
| 5 | | | | | |
| 6 | | | | | |
| 7 | | | | | |
| 8 | | | | | |
| 9 | | | | | |
| 10 | | | | | |

| | | | |
|---|---|---|---|
| HR08 | **I have recorded that you were XX persons living together on October 6. Is that correct?** | 1: yes<br>2: no | *àHR01 and revise* |
| HR09 | **I have recorded that xx,yy,zzz (read list of persons, or say "no one has left your household") have left your household. Is that correct?** | 1: yes<br>2: no | *àHR01 and revise* |
| HR10 | **I have recorded that xx,yy,zzz (read list of persons or say "no one has died in your household") have died since October 6,2023. Is that correct?** | 1: yes<br>2: no | *àHR01 and revise* |
| HR11 | **I have recorded that xx,yy,zzz (read list of persons or say "no one has gone missing in your household") have gone** | 1: yes<br>2: no | *àHR01 and revise* |

| | | | |
|---|---|---|---|
| | **missing since October 6,2023. Is that correct?** | | |
| HR12 | **Between October 6, 2023 and today were any children born to any woman you have just listed as a household member?** | 1: Yes<br>2: No | <br>*àEnd interview (ID08)* |

**HR13: Has any member of your family, other than those who lived with you in the same house on October 6, been killed during the current war that started on October 7?** 1) Yes 2) No 99) Do not know

#### Section 4: Children

***Please list the children that have been born to any woman of the household on or after October 6, 2023***

| BR01: **Serial number of Birth** | BR02: **Name of child** (The name will only be used for reference during the interview and will not be included in the data file) | BR03: **Who is the mother?** (serial number from HR01) | BR04: **Date of birth** Yy,mm,dd (Depending on CAPI software, record in three variables, or use date field) | BR05: Gender<br>1: Male<br>2: female | BR07: **Is …. still living in the household or has he/she moved, died, or is missing**<br>1. Still resident à Next person<br>2. Left Gaza à Next person<br>3. Moved within Gaza à Next person<br>4. Dead<br>5. Missing à Next person | BR08: **How old was … when he/she died?**<br>Record in BR08A:<br>1: Days<br>2: months<br>3: Date (yy,mm,dd)<br>4: Not known<br>Use BR08_1,BR08_2, BR08_3 for recording | BR09: **What was the reason for the death?**<br>1. Disease, pre-existing conditions etc<br>1. Disease, pre-existing conditions etc, not receiving medical aid<br>2. Accident<br>3. Killed in/because of fighting<br>4. Do not know |
|---|---|---|---|---|---|---|---|
| 1 | | | | | | | |
| 2 | | | | | | | |

| |
|---|
| 3 |
| 4 |
| 5 |

| | | | |
|---|---|---|---|
| BR10 | **I have recorded that there were XX births to members of your household since October 6. Is that correct?** | 1: yes<br>2: no | *àBR01 and revise* |

#### Section 5: End of interview

***Thank you for participating in this survey***. (Any other messages here?)

| | | | |
|---|---|---|---|
| ID08 | **Time of interview end** | To be filled in by CAPI | __:__ |
| ID09 | **Duration of interview** | To be filled in by CAPI | |
| ID10 | **Supervisor present during interview** | 1: yes<br>2:no | |

##### *Notes for CAPI*

ID01,02,03,07 should be loaded/verified from a table allocating interviewers to locations.

HR08: Compute from roster

HR09-HR11: Get the list of relevant persons' names from the roster.

HR02, BR02: Names should not be stored in data file transmitted from the device

HR03: Should be within range [0,110]

HR07 should be in range [HR03, HR03+2]

BR03 must refer to a woman 15+ in the HR section. If not, interviewers must ask for clarification and, if needed, revise HR section.

BR08 must be less than date of Interview (ID04) – date of birth (BR04)

BR04 Should be valid date in range [2023.10.7, ID04]

##### *Field Check Tables*

The following field check tables should be produced on a daily basis

a. Interviews per day and cumulative number of interviews.

b. Locations visited

c. Household size distribution

d. Average household size by interviewer, location, region of the Gaza strip and total

e. Number of deaths recorded per household by interviewer, location, region of the Gaza strip and total

f. Gender and age distribution of the household population

g. Gender distribution of the household population by interviewer, total

h. Number of children by interviewer, total

i. Cause of death by the interviewer, total

j. Duration of interview by interviewer, total

k. Distribution and average of time between interviews by interviewer, total

l. Percentage of ages with the last digit being 0 or 5 by interviewer, total
